# Fractional dosing to improve vaccination coverage, vaccine hesitancy, and cost-effectiveness in Africa: a systematic review

**DOI:** 10.1101/2023.08.28.23294558

**Authors:** Abdourahamane Yacouba, Souleymane Brah, Maman Daou, Abdoul-Kader Andia, Alkassoum Ibrahim, Lamine Mahaman Moustapha, Daouda Alhousseini, Mahamadou Doutchi, Ousmane Guindo, Issaka Soumana, Matthew E. Coldiron, Saidou Mamadou, Eric Adehossi, Rebecca F. Grais

## Abstract

**Background:** The major challenges of vaccination programs are notably coverage in the target population, vaccine hesitancy, and cost-effectiveness. A vaccination strategy with fractional doses is a dose-saving strategy to address current vaccine shortages. Herein, we aimed to review the literature on administering fractional vaccine doses in Africa.

**Methods:** A methodical search of PubMed was conducted to identify articles published up till March 31, 2023. Peer-reviewed studies were selected for inclusion if they focused on studies that described the use of fractional doses of vaccines and were conducted in any of the 54 African countries.

**Results:** Findings from eleven eligible studies were analyzed. Studies were from the Democratic Republic of the Congo, Gambia, Ghana, Kenya, South Africa, and Uganda. They covered five vaccines including the yellow fever vaccine (n=3; 27.3%), inactivated poliovirus vaccine (n=3; 27.3%), meningococcal A/C/Y/W135 vaccine (n=2; 18.2%), *Haemophilus influenzae* type b vaccine (n=2; 18.2%), and malaria vaccine (n=1; 9.1%). Fractionated doses used most often consist of one-fifth of the standard dose (n=8; 72.7%). Regarding immunogenicity/efficacity, eight of ten studies that addressed immunogenicity suggest that immune responses to the fractional dose vaccines were comparable to that of the standard dose vaccines and resulted in higher antibody titers. Regarding safety, all of the eight studies that addressed the safety of fractional doses in Africa, suggest that safety and tolerability data of fractional dosing were favorable compared to full dose regimen.

**Conclusion:** Fractional dosing may be considered to address the availability and acceptability of certain vaccines while maintaining protection. This strategy also has the potential advantage of reducing the cost of vaccination programs, vaccine antigen overload, and vaccine side effects.

## Introduction

Vaccination is acknowledged as the most effective and cost-effective means of preventing and reducing the severity of infections [1,2]. Vaccination may further reduce the magnitude epidemics and their consequences on hospital capacity by reducing case loads thereby preserving the capacity of healthcare institutions to manage all acute and chronic pathologies [3,4]. According to the World Health Organization (WHO), vaccination could prevent 3.5 to 5 million deaths each year worldwide [5]. These numbers could increase by 1.5 million if global immunization coverage improves in all countries [5]. The expected effect of the implementation of a vaccination campaign is to obtain vaccination coverage allowing for the reduction in the circulation of the identified infectious agent in the population while protecting individuals [6,7]. To achieve these goals, and derive maximum benefit, vaccination coverage must reach certain critical levels for different pathogens if herd immunity is a goal of the vaccination program [8].

According to WHO, reluctance or refusal to vaccinate is complex and does not depend on a simple set of individual factors [9]. It is important to understand these concerns and develop strategies to facilitate the acceptance of vaccination programs. In addition to the problem of vaccine refusal, some countries will face vaccine stock shortages with the global outbreak of epidemics straining the global vaccine supply [10,11].

Fractional dosing is a dose-saving approach under consideration to address vaccine stockouts [12]. For example, in yellow fever, fractional dosing relies on the fact that the minimum amount of virus required to achieve a protective titer of neutralizing antibody is 1000[IU [13], whereas the standard vaccine dose typically contains ≥ 10,000 IU of the virus. Fractional doses contain one-third or one-fifth or one-tenth or one-fiftieth of the standard dose. In practice, when a full-dose vaccine vial is used to deliver fractional doses, the number of doses in the vial is increased threefold or fivefold or tenfold: e.g., a vial containing 5 or 10 full doses becomes a vial of 25 or 50 fractional doses, respectively.

The objective of this review was to analyze the literature on the strategy of administering fractional doses of vaccines in Africa, focusing on efficacy, immunogenicity, and safety. Although efforts are currently underway to increase the possibility of vaccine manufacturing on the African continent, fractional dosing strategies may also be needed in the future and potentially offer other benefits.

## Materials and Methods

### Literature search

We searched PubMed to identify articles related to fractional doses of vaccines in humans in Africa. Two authors (AY and BS) independently performed the literature search. Keywords used for the search were “fractional doses” “fractional dosing”, “Vaccines”, “Africa”, and specific names of all African countries. The detailed search strategy can be found in S1 Appendix file. A manual search for additional studies was performed using references cited in original study articles and reviews. Additionally, studies were retrieved by searches of the WHO vaccines and immunization Infobase. To avoid the inclusion of duplicate studies, publications were cross-referenced considering the place and time period of reported studies.

### Study selection

Publications identified were considered up to March 31, 2023. Literature published either in English or French was considered. Studies identified in the initial search were first screened by title and abstract and retained if they met the predefined inclusion criteria, as follows: (i) original article published in a peer-reviewed journal, (ii) studies that described the use of fractional doses of vaccines, and (iii) studies conducted in any of the 54 Africa countries. Expert opinions, review articles, modeling study, animal models, and protocols were excluded. Publications were reviewed by two independent researchers (AY and BS) to determine whether they met inclusion/exclusion criteria and if disagreement was arbitrated by MD.

### Data extraction

Information extraction was done using Microsoft Excel 2013 on a predesigned database, developed for the purposes of this review. Data extracted included article information (first author, year of publication, and country), study design (type of study, sample size, age group), vaccines used, type of fractionated doses, and main findings. Additionally, to address the immunogenicity of fractional doses of vaccines, a complementary information including seroconversion percentage and geometric mean concentrations (GMC) were extracted.

### Data synthesis and analysis

A literature synthesis was performed based on the information gathered from the analyzed publications. Qualitative data were presented as effective and proportion. Statistical analyses and visualization were performed using R-software version 4.0.4. Summary figures describing the main findings were made using the Biorender application (https://app.biorender.com).

## Results

### Literature search

A total of 183 potentially relevant articles on fractional dose vaccine strategy in Africa were identified in the initial electronic search. The titles and/or abstracts of 183 studies were screened for relevance. Of the 183 studies screened, 152 studies were excluded and the remaining 31 studies were retrieved for full text review. Ultimately, 11 studies were included in this review (Fig 1).

**Fig 1.**
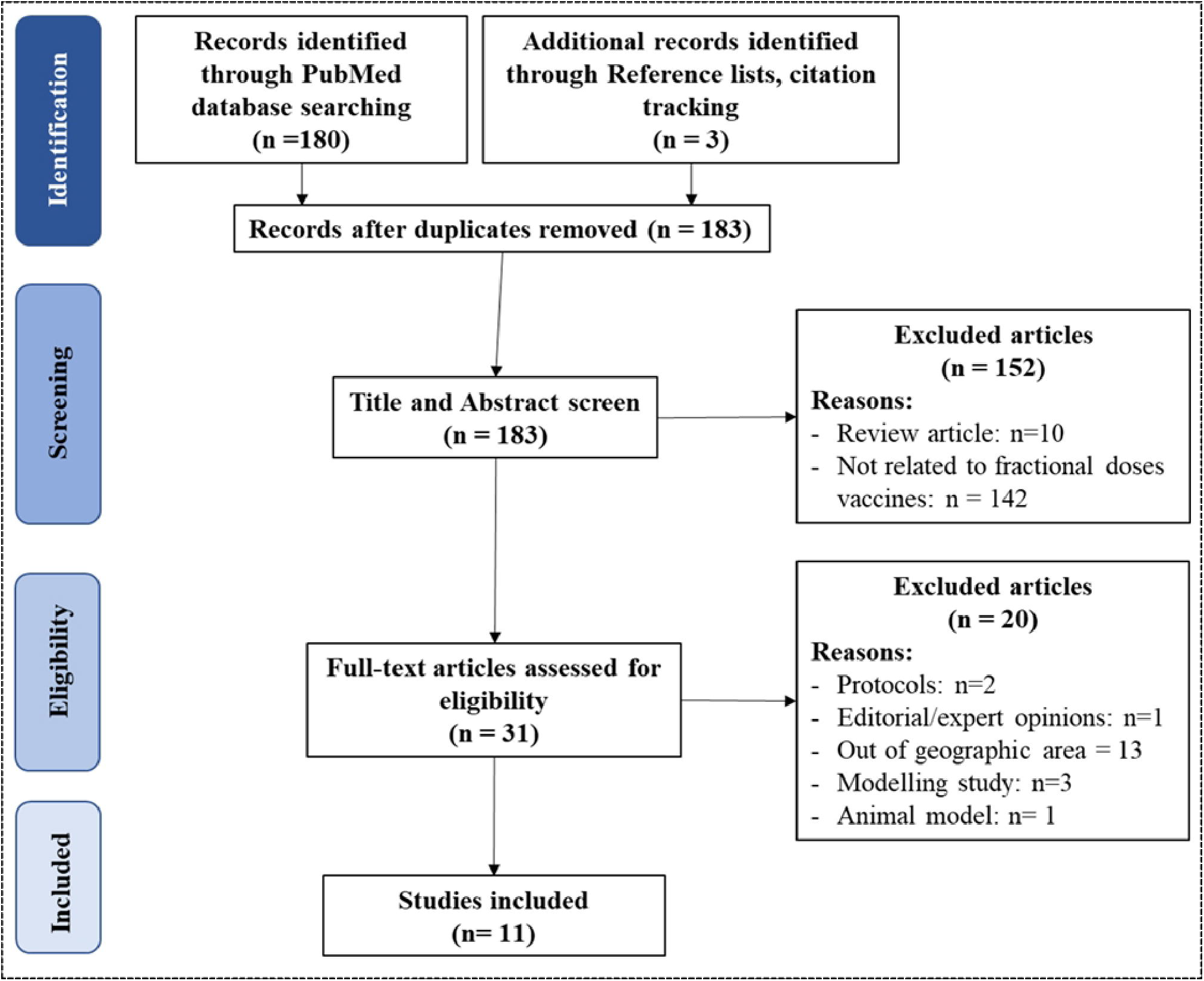
Flow diagram of the selection process of the included studies.

### Data and study characteristics

Of the 11 studies included, two (18.2 %) were from Democratic Republic of the Congo [16,17], two (18.2 %) from Gambia [18,19], two (18.2 %) from South Africa [20,21], three (20.0 %) from Uganda [22–24], including one (9.1 %) performed in two countries, Uganda and Kenya [24] (S1a Fig).

Vaccines fractionated in Africa were mostly yellow fever vaccine [16,17,24], (n=3; 27.3%) and inactivated poliovirus vaccine [18,19,25], (n=3; 27.3%), followed by meningococcal A/C/Y/W135 vaccine [22,23], (n=2; 18.2%) (Table 1).

**Table 1.** Vaccines fractionated in Africa.

| vaccines | Number | Percentage |
| --- | --- | --- |
| *Haemophilus influenzae* type b vaccine | 2 | 18.2 |
| Inactivated Poliovirus vaccine | 3 | 27.3 |
| Malaria vaccine | 1 | 09.1 |
| Meningococcal A/C/Y/W135 vaccine | 2 | 18.2 |
| Yellow fever vaccine | 3 | 27.3 |

|  | Total | 11 | 100.0 |
| --- | --- | --- | --- |

Fractionated doses used in Africa consist most often of one-fifth (1/5) of standard dose (n=8; 72,7%) [16–19,22–24,26]. Other dilution used as fractional doses included 1/10 [21–23] and half (1/2) [25] of the full dose (Table 2). The result of distribution by time indicates that the literature on fractional dosing strategy in Africa was first published in 2002 in South Africa [21] (S1b Fig). Studies consisted of randomized and controlled trials [17–26] (n=10; 90,9%) and community-based pharmacovigilance [16] (n=1; 9.1%). Of the randomized trial, half (5/10) were non-inferiority trial (Table 2).

**Table 2:** Mains findings related to fractional doses vaccination in Africa.

| Reference | country | vaccines | Type of study | Participants number | Intervention | Immunogenicity |  |  | Long term protection | Safety |
| --- | --- | --- | --- | --- | --- | --- | --- | --- | --- | --- |
|  |  |  |  |  |  | Seroconversion, % (CI 95%) | Geometric mean concentrations (GMC) in Ig/ml (CI 95%) | Findings |  |  |
| Casey et al. 2019 [17] | Democratic Republic of the Congo | Yellow fever | randomized controlled trial | 764 | 1/5 of standard dose | **In one month:** 98 % [96–99];<br>**In one year:** 96 % [94–98] | **In one month:** 1340 (1117–1607);<br>**In one year:** 143 (123–166) | The immunologic response to a fractional dose of the 17DD yellow fever vaccine was appropriate for a response to a yellow fever outbreak among children 2 years of age or older and among nonpregnant adults | Not reported | Not reported |
| Barnes et al. 2011 [23] | Uganda | <u>Menomune</u> from Sanofi Pasteur, a tetravalent <u>A/CY/W135</u> polysaccharide vaccine | randomized controlled trial | 115 | 1/5 and 1/10 of full dose | Not reported | 1 month after the first dose:<br>**1/10 of full dose** 43.3 (36.5–51.4)<br>**1/5 of full dose** 44.6 (38.4–51.8) | Fractional doses are able to elicit antibodies of as good avidity against serogroup A polysaccharide as a full dose | Not reported | Not reported |
| Bashorun et al. 2022 [19] | Gambia | Inactivated poliovirus vaccine | pragmatic, open-label, non-inferiority trial | 3170 | 1/5 of full dose | Polyovirus 1: 88.3% (80.6–93.1)<br>Polyovirus 2: 100.0% (93.1–100.0)<br>Polyovirus 3: 81.9% (76.5–86.1) | Polyovirus 1: $\geq 1448$ (1448–1448)<br>Polyovirus 2: $\geq 1448$ (1448–1448)<br>Polyovirus 3: $\geq 1448$ (1448–1448) | Non-inferiority was demonstrated between the full dose and fractional doses when administered with an BCG needle and syringe, a disposable syringe jet injector, and an intradermal adaptor | Not reported | Safety and tolerability data were favorable |
| Clarke et al. 2016 [18] | Gambia | Inactivated poliovirus vaccine | randomized, non-inferiority trial | 182 | 1/5 of full dose (0.1 ml) | Polyovirus 1: 86.4% (66.7–95.3)<br>Polyovirus 2: 100.0% (67.6–100.0)<br>Polyovirus 3: 90.4% (79.4–95.8)<br>Measle: | Not reported | fractional doses did not achieve non-inferiority compared with full dose | Not reported | Safety and tolerability data were favorable |
| Guerin et al. 2008 [22] | Uganda | A/C/Y/W135 polysaccharide meningococcal vaccine | Randomized Non-Inferiority Controlled Trial | 750 | 1/10, 1/5 of full dose | **PerProtocol (PP) population1/5 of full dose**<br>Serogroup A : 77.5 (72.0–83.0)<br>Serogroup W135 : 94.6 (91.6–97.6)<br>Serogroup C : 80.4 (75.2–85.6)<br>Serogroup Y : 82.4 (77.4–87.4)<br>**1/10 of full dose**<br>Serogroup A : 69.4 (63.5–75.3)<br>Serogroup W135 : 95.6 (93.0–98.2)<br>Serogroup C : 76.6 (71.1–82.1)<br>Serogroup Y : 83.9 (79.2–88.6) | **PerProtocol (PP) population1/5 of full dose**<br>Serogroup A : 2054.4 (1612.2–2618.0)<br>Serogroup W135 : 2041.6 (1582.2–2634.5)<br>Serogroup C : 467.3 (328.0–665.8)<br>Serogroup Y : 768.3 (524.4–1125.6)<br>**1/10 of full dose**<br>Serogroup A : 1369.3 (1083.3–1730.7)<br>Serogroup W135 : 2426.3 (1979.7–2973.7)<br>Serogroup C : 396.3 (274.6–572.0)<br>Serogroup Y : 816.8 (565.1–1180.8) | Non-inferiority was demonstrated between the full dose and fractional doses against serogroups W135, Y and A. Fractional doses did not achieve non-inferiority compared with full dose for serogroup C. | Not reported | Safety and tolerability data were favorable |
| de Deus et al. 2020 [25] | Mozambique | monovalent type 2 oral poliovirus vaccine (mOPV2) | randomized, controlled, open-label, noninferiority trial | 378 | ½ of the full dose | 53.6% (44.9%–62.1%) | Not reported | Fractional doses did not achieve non-inferiority compared with full dose | Not reported | Not reported |
| Huebner et al. 2004 [20] | South Africa | Haemophilus influenzae type b vaccine conjugated either to diphtheria CRM197 (Chiron, Sienna, Italy) or tetanus toxoid (Berna Biotech Ltd., Berne, Switzerland) | randomized trial | 168 | 1/16; 1/8; 1/4; 1/2 of full dose | **1/8 of full dose**<br>100 % (95–100) | **1/8 of full dose**<br>6.33 (4.21–9.50) | vaccines of $\geq 1.25$ $\mu\text{g}$ (1/8 of full dose) may be sufficient to stimulate an immune response that offers both short and longer-term protection from invasive Hib disease | Offer longer-term protection from invasive Hib disease | Safety and tolerability data were favorable |
| Nicol et al. 2002 [21] | South Africa | Haemophilus influenzae type b polysaccharide-tetanus toxoid conjugate (PRP-T) vaccine | randomized trial | 168 | 1/10 of full dose | 94% | Not reported | The 1/10 dose of PRP-T was as immunogenic as the full dose. | Not reported | Safety and tolerability data were favorable |
| Juan-Giner et al. 2021 [24] | Kenya and Uganda | Yellow fever: 17DD Bio-Manguinhos-Fiocruz, 17D-213 Chumakov Institute of Poliomyelitis and Viral Encephalitis, 17D-204 Institut Pasteur Dakar, 17D-204 Sanofi Pasteur | randomized, double-blind, non-inferiority trial | 960 | 1/5 of full dose | **Day 28 17DD Bio-Manguinhos-Fiocruz:** 100.0 % (96.7 to 100.0); **17D-213 Chumakov Institute of Poliomyelitis and Viral Encephalitis:** 99.1% (95.1 to 100.0); **17D-204 Institut Pasteur Dakar:** 100.0 % (96.8 to 100.0); **17D-204 Sanofi Pasteur:** 100.0 % (96.8 to 100.0) | **Day 28 17DD Bio-Manguinhos-Fiocruz:** 3939 (2812, 5516); **17D-213 Chumakov Institute of Poliomyelitis and Viral Encephalitis:** 5874 (4162 to 8289); **17D-204 Institut Pasteur Dakar:** 4279 (3182 to 5753); **17D-204 Sanofi Pasteur:** 5545 (4106, 7488) | non-inferior to the standard dose in inducing seroconversion 28 days after vaccination | Not reported | Safety and tolerability data were favorable |
| Samuels et al. 2022 [26] | Ghana | Malaria vaccine | randomized controlled trial | 2157 | 1/5 of full dose | Not reported | Not reported | 1/5 of full dose regimen does not affect protective efficacy over | Not reported | Safety and tolerability data were favorable |
| Nzolo et al. 2018 [16] | Democratic Republic of the Congo | 17DD Yellow Fever Vaccine | Community-based pharmacovigilance | 4020 | 1/5 of full dose | Not reported | Not reported | Not reported | Not reported | Safety and tolerability data were favorable |

### Advantages of fractional doses strategy

Evidence synthesis on the advantages of fractional dosing strategy highlighted that in addition to the reduction of vaccine antigen overload, reduction of vaccine cost, reduction of stock-outs, and long-term durable protection, fractional doses retain their immunogenicity and have good safety and efficacy profiles (Fig 2).

**Fig 2.**
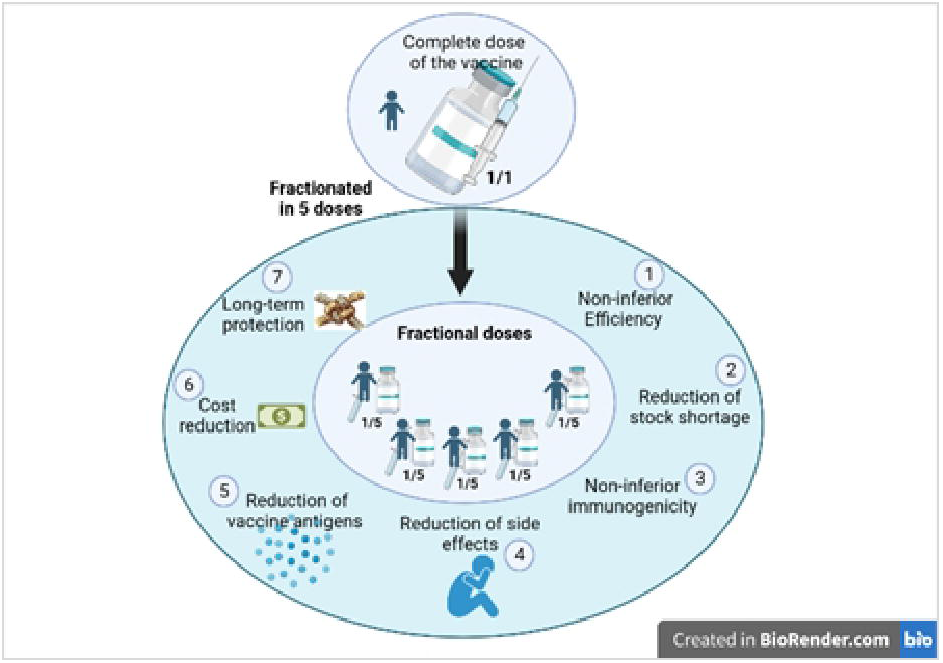
Advantages of vaccination using fractional dosing strategy.

#### Immunogenicity and efficiency of fractional dosing strategy

Ten of eleven studies [17–26] addressed immunogenicity of fractional doses strategy in Africa. Of the ten, two studies [18,25] did not establish non-inferiority of the fractional dose strategy compared with a full dose (Table 2). In one study [26], authors reported that 1/5 of the full dose was as effective as the standard dose of the malaria vaccine (Table 2). Evidence from eight studies [17,19–24] suggests that the fractional dosing strategy could yield high immune responses, comparable to those for standard doses of the same vaccine (Table 2). These authors support the use of fractional dose vaccination for outbreak control [17,19–24].

#### Safety of fractional dosing strategy

Eight of eleven (72.7%) studies [16,18–22,24,26] addressed the safety of fractional doses strategy in Africa. All of them suggest that safety and tolerability data of fractional dosing were favorable compared to the full dose regimen.

## Discussion

The use of a fractional dosing strategy is a well-known strategy [12,27,28]. Our study demonstrated that a fractional dosing strategy may be as sufficient to stimulate an immune as a standard dose of vaccine. This means that, for yellow fever, inactivated poliovirus, A/C/Y/W135 polysaccharide meningococcal, and *Haemophilus influenzae* type b vaccines, immunogenicity data suggest that 1/5 or 1/10 doses could be almost as efficacious as standard dose used of the same vaccines. Authors reported that the 1/5 of standard dose of the 17DD Yellow fever vaccines was effective at inducing seroconversion in 98% of the participants who were seronegative at baseline, which is similar between gender [17]. In a recent study, seroconversion rates at four to five weeks following vaccination were similar between participants who received standard doses and participants who received fractional doses containing one-third, one-fifth, one-tenth, and one-fiftieth of the standard dose [12]. Among the immunological mechanisms proposed to explain this phenomenon, one can cite the fact that dose fractionation favors the persistence of antigen-specific B lymphocytes [29]. The increase in somatic hypermutation and antibody avidity [30,31] has high responses to immunoglobulin G class 4 [32] and higher neutralizing antibody titers [33]. Additionally, fractional doses may also have less side effects [34], vaccine antigen overload, cost, and stock-outs [35,36,17] than standard dose. Vaccine shortages and stockouts during outbreaks happen frequently in underdeveloped countries, and occasionally in developed countries [37–40]. In a country, the shortage/stockout events can occur at the national and at subnational level. The most frequently reported causes vaccine shortages/stockouts in Africa include global shortage, disease outbreaks, poor stock management, poor supply chain structure, delays in deliveries, lack of trained health personnel, and lack of resource to purchase vaccine [41,42]. Typically, to address vaccine shortages/stockouts, actions undertaken by African countries were to purchase additional doses of the vaccine from other supplier/manufacturer, the use of available stockpiles of vaccine, to redistribute stock doses among regions and facilities, to import vaccine from another country, or to contact WHO (or other institutions) for technical assistance. On top of that, using lower doses of vaccine is one such action [17]. Currently, the fractional-dose strategy is emerging as an alternative option for urgently stretching restricted vaccines supplies [10,11,35]. WHO supports the use of fractional doses strategy for yellow fever as part of an emergency response to an epidemic if the shortage of full doses exceeds the capacity of the global stockpile [36]. Many countries in developed and underdeveloped countries have experimented with the use of fractional doses to address vaccine shortages [10,11,35].

Studies showed that one of the most serious concerns influencing vaccination acceptance are about side effects [43–45]. Individuals or parents were concerned about potential side effects of vaccines [46]. According to Saied et al. [43] in Egypt, 96.8% of the participants of their study had concerns regarding the vaccine’s adverse effects [43]. In a community based-pharmacovigilance, fractional dosing of 17DD Yellow fever vaccine has a good tolerability and safety profile, which is almost similar between females and males [16]. Fractional doses of the same vaccine may be superior if they offer comparable efficacy with lesser side effects. If efficacy is comparable to that of standard doses, and side effects are lesser, fractional doses might be superior to current doses in terms of individual benefit–risk profile.

### Fractional doses strategy to address vaccination coverage in Africa

Childhood vaccination is one of the fundamental strategies for achieving goal three of the Sustainable Development Goals (SDGs), which is to reduce under-five mortality to less than 25/1000 live births by 2030 [47]. In Africa, vaccination coverage is lagging behind the 90% target set in the regional strategic plan for vaccination 2014-2020 [8]. As an example, vaccination coverage remained low in 2017 for DTP3 (72%), PCV3 (68%), Hib3 (72%), MCV1 (70%), and RCV1 (26%) [48]. Many barriers exist to achieving good vaccination coverage in Africa, including factors associated with vaccine hesitancy.

### Fractional doses strategy to address vaccine hesitancy

There are several possible reasons for vaccine refusal (Fig 3). The main factors are religion, parental distrust, dissemination of misinformation, and anti-vaccine movements. According to WHO, vaccine hesitancy or refusal is the 8th most prevalent health threat after pollution and climate, non-communicable diseases, pandemic influenza, vulnerable countries, antimicrobial resistance, Ebola and high-risk pathogens, and primary health care [49,50]. Vaccination denial dates back to the first vaccine administered in humans, the smallpox vaccine [51].

**Fig 3.**
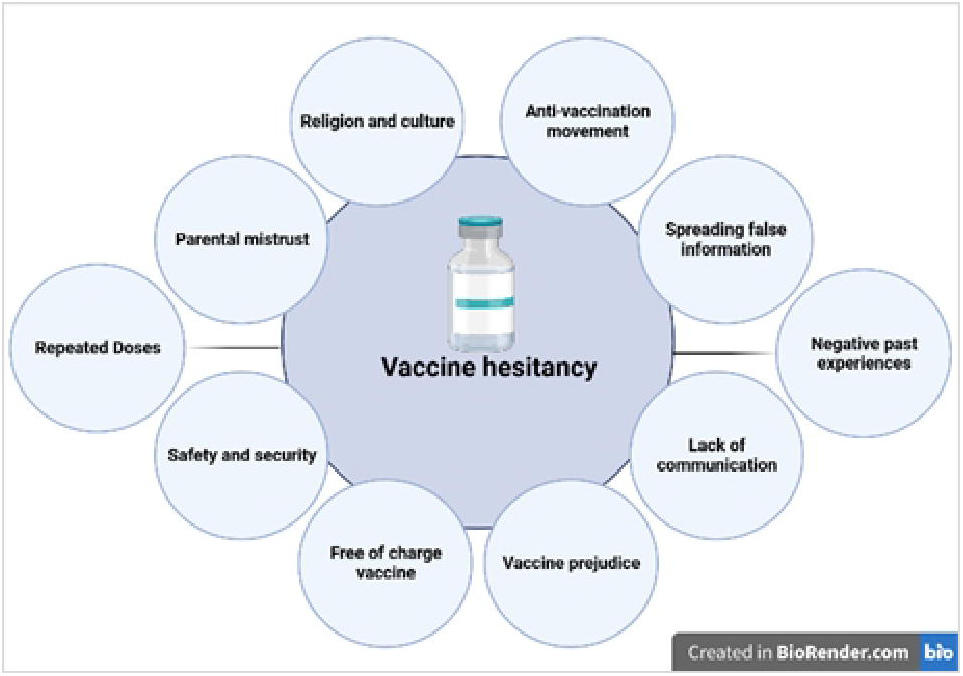
Main factors behind vaccine hesitancy.

Parents’ decisions about whether or not to vaccinate their children are complex and multidimensional. However, the main parent-specific factor most often involved in their experience with vaccination (side effects) [52–56]. In addition to keeping parents away from the vaccine, the presence of side effects could amplify the spread of misinformation about the vaccine anti-vaccine movements. Fractional dose strategies may resolve this issue by reducing the side effects of the vaccines compared to full dose [12,24,57–61].

The rise of well-organized anti-vaccine movements is currently a serious threat to vaccination programs in both developed and developing countries. The main concerns put forward by the anti-vaccine community were antigenic overload [62,63], autoimmunity [64–68]. By reducing vaccine antigen overload compared to full dose regimen, fractionated doses strategy may address this issue.

Cost-effectiveness analyses are critical to planning vaccination campaign financing decisions. It is a measure of the amount of money needed to fund an vaccination project and achieve the previously defined goal. Fractional doses vaccines could be an economically viable vaccination strategy compared to full-dose vaccination or no vaccination [69]. Additionally, fractional doses vaccines strategy could save a large number of lives, and for mitigated the public health costs of resurgences after vaccination [70].

### Strengths and Limitations

To the best of our knowledge, this study is the first to review the vaccination strategy with fractional doses strategy in Africa, a topic with potential significant public health impact. However, it is limited by the relatively few publications on fractional dosing strategy in Africa that may impact generalizability. Another flaw in the current systematic review that may impact generalizability is that the majority of the studies are from the Southern Africa. There is, therefore a need for a continuous update, especially on the impact of fractional dosing strategy on vaccine hesitancy, as new evidence emerges. Another shortcoming is that meta-analysis is not performed, which might impact power to study the factors associated to fractional dose strategy. Even then, this study synthesized the current state and should help inform policy decision-making and research needs for fractional dosing strategy in Africa.

## Conclusion

In Africa, despite the progress made in recent years in the various immunization programs, vaccination coverage is far from reaching the 90% target. Various complex factors are at the origin of vaccine hesitancy. The use of fractional doses vaccine strategy could be a solution to improve the availability and acceptability of vaccines while saving costs, reducing side effects, and maintaining efficacy, immunogenicity, and long-term protection.

## Supporting information

**S1 Fig. Country (a) and time distribution (b) of studies on fractional doses vaccines.**

## Supporting information

Prisma checklist

## Data Availability

The authors confirm that the data supporting the findings of this study are available within the article [and/or] its supplementary materials.

## Notes

### Competing Interest Statement

The authors have declared no competing interest.

