## Supplementary material for "Fractional dosing to improve vaccination coverage, vaccine hesitancy, and cost-effectiveness in Africa: a systematic review": S1 appendix

### **Search strategy**

The request consists of combinations of the following keywords : “fractional doses” “fractional dosing”, “Vaccines”, “Africa”, and specific names of all African countries. The search fields were connected with “AND“ in order to ensure that at least one of the terms can be found in the results. All terms in one search field were linked with the conjunction “OR“.

PubMed (March 31, 2023) (180)

((fractional dos*[All Fields]) OR (fractionated dos*[All Fields])) AND "vaccines"[MeSH Terms] AND ("africa"[MeSH Terms] OR "africa"[All Fields] OR "africa s"[All Fields] OR "africas"[All Fields]) AND ((Africa*[All Fields]) OR "Comoros"[Mesh] OR "Djibouti"[Mesh] OR "Madagascar"[Mesh] OR "Malawi"[Mesh] OR "Seychelles"[Mesh] OR "Cameroon"[Mesh] OR "Central African Republic"[Mesh] OR "Chad"[Mesh] OR "Congo"[Mesh] OR "Equatorial Guinea"[Mesh] OR "Atlantic Islands"[Mesh] OR (Gabon*[All Fields]) OR "Morocco"[Mesh] OR "South Sudan"[Mesh] OR "Sudan"[Mesh] OR "Botswana"[Mesh] OR "Lesotho"[Mesh] OR "Swaziland"[Mesh] OR "Benin"[Mesh] OR "Burkina Faso"[Mesh] OR "Cape Verde"[Mesh] OR "Ghana"[Mesh] OR "Guinea"[Mesh] OR "Guinea-Bissau"[Mesh] OR "Mauritania"[Mesh] OR "Niger"[Mesh] OR "Senegal"[Mesh] OR "Sierra Leone"[Mesh] OR "Togo"[Mesh] OR (Burundi*[All Fields]) OR (eritrea*[All Fields]) OR (ethiopia*[All Fields]) OR (kenya*[All Fields]) OR (mozambique*[All Fields]) OR (rwanda*[All Fields]) OR (somalia*[All Fields]) OR (tanzania*[All Fields]) OR (uganda*[All Fields]) OR (zambia*[All Fields]) OR (zimbabwe*[All Fields]) OR (angola*[All Fields]) OR (algeria*[All Fields]) OR (egypt*[All Fields]) OR (tunisia*[All Fields]) OR (namibia*[All Fields]) OR (south africa*[All Fields]) OR (gambia*[All Fields]) OR (liberia*[All Fields]) OR (mali*[All Fields]) OR (Nigeria*[All Fields])) AND Humans[Mesh]
