## Supplementary figures and images for "Fractional dosing to improve vaccination coverage, vaccine hesitancy, and cost-effectiveness in Africa: a systematic review"

### S1 Fig

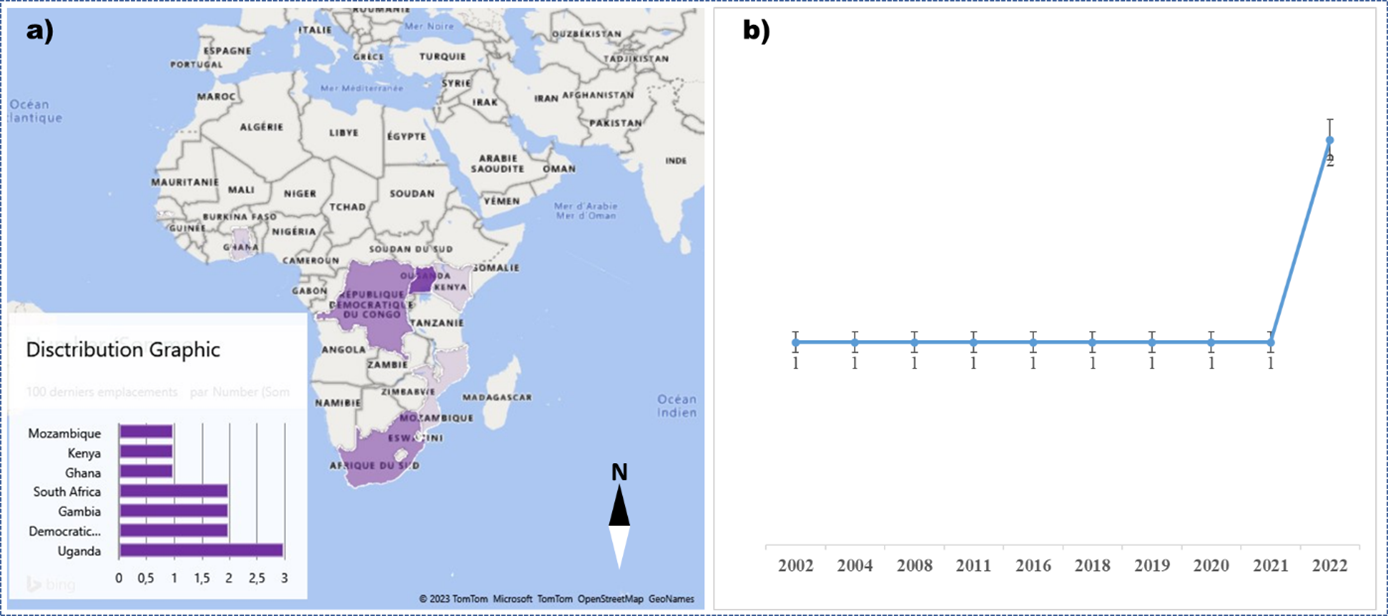
